# Test-Retest Reliability of Artificial Intelligence-Enhanced Electrocardiography: A Multi-Center Study

**DOI:** 10.1101/2025.11.04.25339526

**Authors:** Lovedeep S Dhingra, Philip M Croon, Bruno Batinica, Evangelos K Oikonomou, Rohan Khera

**Affiliations:** Cardiovascular Data Science (CarDS) Lab, Yale School of Medicine, New Haven, CT, USA; Section of Cardiovascular Medicine, Department of Internal Medicine, Yale School of Medicine, New Haven, CT, USA; Department of Cardiology, Amsterdam Cardiovascular Sciences, Amsterdam University Medical Center, University of Amsterdam, Amsterdam, The Netherlands; Center for Outcomes Research and Evaluation (CORE), Yale New Haven Hospital, New Haven, CT, USA; Department of Biomedical Informatics and Data Science, Yale School of Medicine, New Haven, CT, USA; Section of Health Informatics, Department of Biostatistics, Yale School of Public Health, New Haven, CT, USA

**Keywords:** Artificial Intelligence, Implementation Science, Deep Learning, Structural Heart Disease, Test-Retest Reliability, Electrocardiograms, Echocardiography

## Abstract

**Background:** Artificial intelligence-enhanced electrocardiography (AI-ECG) enables structural heart disease (SHD) detection. However, its utility as a clinical assay requires consistency of outputs when repeated under similar conditions.

**Objectives:** To characterize test-retest reliability of contemporary AI-ECG models across diverse health systems, identify factors associated with discordance, and determine the predictive significance of screen status changes.

**Methods:** We identified ECG pairs recorded 1-30 days apart in the same individual at the Yale-New Haven Health System (YNHHS), Massachusetts General Hospital (MGH), and Emory University Hospital (EUH). We evaluated internally developed ECG signal- and image-based SHD models, and the EchoNext-Mini model, including disease-specific components and ensemble composites. Reliability was quantified with concordance correlation coefficients (CCCs) and categorical concordance percentages. In patients without prevalent heart failure (HF) with serial ECGs 30-90 days apart, we evaluated association of screen status discordance with new-onset HF risk.

**Results:** We included 731,466 ECG pairs (median interval 5-6 days). At YNHHS, disease-specific model CCCs ranged from 0.77-0.86 across the signal- and image-based model families and 0.50-0.97 for EchoNext-Mini output nodes. Composite SHD models had CCCs of 0.90 (signal-based), 0.90 (image-based), and 0.81 (EchoNext-Mini). The image-based ensemble model achieved categorical screen-status concordance of 87-89% across 3 health systems (versus 80-84% for disease-specific models). Younger age (< 65 years) was the dominant correlate of discordance (OR 1.69 [95% CI, 1.65-1.74]), along with higher odds for inpatient ECGs (ORs 1.30-1.41). Among 65,838 individuals in the predictive cohort, a negative-to-positive change in screen status was associated with elevated risk of new-onset HF (adjusted hazard ratio 1.67-2.37 across sites).

**Conclusions:** Contemporary AI-ECG models had high test-retest reliability, with composite ensembles showing the greatest stability. Serial changes in AI-ECG screen status carry predictive information for new-onset HF.

**CONDENSED ABSTRACT:** Clinical utility of artificial intelligence-enhanced electrocardiography (AI-ECG) requires consistency when repeated under similar conditions. In 731,466 ECG pairs from 3 health systems, AI-ECG models for structural heart disease (SHD) had high test-retest reliability, with composite SHD models showing the greatest stability (concordance correlation coefficient 0.87-0.90). Signal- and image-based models had comparable reliability. Younger age and inpatient acquisition were the primary correlates of screen-status discordance. Among 65,838 individuals at risk, a negative-to-positive change in screen status independently predicted new-onset heart failure (adjusted hazard ratio 1.67-2.37 across sites). Test-retest reliability and serial screen-status changes present meaningful clinical benchmarks for AI-ECG implementation.

## BACKGROUND

The application of artificial intelligence to electrocardiograms (AI-ECG) has defined a new paradigm for cardiovascular diagnostics.^1,2^ Deep learning models can detect structural heart diseases (SHDs) including left ventricular systolic dysfunction (LVSD), valvular heart disease, and hypertrophic cardiomyopathy from 12-lead ECGs with strong discrimination across clinical and community-based settings.^3–17^ However, translating this promise of AI-ECG into clinical adoption requires more than high disease discrimination alone.^18–22^ As clinical assays that may alter the course of care, AI-ECG performance must be evaluated for reliability when deployed on repeat measures in the same patient under similar conditions.^21^ Following recent U.S. Food and Drug Administration approvals for AI-ECG tools that detect hidden cardiovascular disorders, such as LVSD, cardiac amyloidosis, and hypertrophic cardiomyopathy, and the reimbursement of AI-ECG for LVSD detection, evaluating retest reliability in real-world settings is a priority.^23^

AI-ECG models have traditionally been validated at a population level using a single ECG per individual, both to ensure generalizability and to avoid over-representing individuals with more frequent testing.^2,5,7–9,24–30^ However, strong aggregate performance in the absence of repeated testing may mask individual-level variability and real-world variation can degrade model performance.^31,32^ Like laboratory assays, AI-ECG provides objective, quantifiable outputs that are independent of inter-reader variability inherent in conventional ECG interpretation. However, while laboratory assays have established coefficients of variation that quantify expected test-retest variability, this remains uncharacterized for AI-ECG. Because AI-ECG models learn high-dimensional representations from complex voltage-time waveforms, they may be susceptible to minor changes in ECG acquisition, such as lead placement, patient positioning, and clinical environment, especially in real-world settings where standardization is limited.^1,2^ Further, while variations in AI-ECG outputs can reflect technical noise, they may also potentially capture clinically-meaningful changes in the underlying cardiovascular physiology. Therefore, the test-retest reliability of AI-ECG models and the clinical significance of output discordance require systematic evaluation.

In this study, we evaluated a range of previously-validated AI-ECG models for SHD detection, using ECG waveform signals or images, across 3 geographically distinct academic US health systems. Our objectives were: (i) to characterize AI-ECG test-retest reliability across different types of models, including individual disease classifiers and models designed to detect a composite of SHDs, across both ECG signal- and image-based implementations; (ii) to identify patient and encounter characteristics associated with model output discordance; and (iii) to evaluate whether discordant AI-ECG results carry predictive significance for new-onset disease.

## METHODS

The Yale Institutional Review Board approved the study protocol and waived the need for informed consent, as this analysis involved the secondary use of previously acquired data.

### Data Sources

We used data from 3 large academic health systems in the United States: the Yale-New Haven Health System (YNHHS), Massachusetts General Hospital (MGH), and Emory University Hospital (EUH).

YNHHS is the largest referral health system in southern New England comprising five hospitals (Yale New Haven Hospital, Bridgeport Hospital, Greenwich Hospital, Lawrence + Memorial Hospital, and Westerly Hospital) and an outpatient provider network. These ECGs were acquired as part of routine clinical care between 2013 and 2025 using the Philips PageWriter and MUSE systems, and were stored in XML format.

MGH is a tertiary academic medical center affiliated with Harvard Medical School in Boston, Massachusetts, and EUH is an academic medical center in Atlanta, Georgia. Both sites collected ECGs during routine clinical care using the MUSE ECG system. ECGs from MGH and EUH were acquired between 1985-2023 and 2012-2023, respectively. These waveforms consisted of 10-second, 12-lead ECGs sampled at 250 Hz (MGH) and 500 Hz (MGH and EUH) and were stored in WFDB format. These data were de-identified following the Safe Harbor method and obtained from the Harvard-Emory ECG Database.^33,34^

### AI-ECG Models

We evaluated 3 families of contemporary AI-ECG models for SHD detection, each containing disease-specific component models and ensemble composites.^5,10^

The first family consists of 7 previously validated ECG signal-based models developed at the Yale New Haven Hospital.^5^ These include 6 convolutional neural network (CNN) models trained to detect individual SHDs, including left ventricular systolic dysfunction (LVSD; left ventricular ejection fraction <40%), severe left ventricular hypertrophy (interventricular septal thickness >15 mm with concurrent diastolic dysfunction), moderate or severe aortic stenosis, moderate or severe aortic regurgitation, moderate or severe mitral regurgitation, and any moderate or severe left-sided valvular disease, and an ensemble composite that integrates these component outputs along with patient age and sex through extreme gradient boosting (XGBoost) to detect a composite of SHDs. The component CNNs use a custom multi-layer convolutional architecture applied to raw 12-lead ECG voltage-time waveforms.

The second family (PRESENT-SHD) includes image-based models, with six analogous disease-specific CNN models and an XGBoost ensemble.^5,11,35^ The image-based models ingest 12-lead ECG images rather than raw waveforms, and were developed using the same disease labels and the 261,228 ECG-echocardiogram development cohort from 93,693 patients at Yale-New Haven Hospital (2015-2023). The image CNN models use an EfficientNet-B3 architecture pretrained with a contrastive self-supervised strategy, capturing generalizable visual representations that were then fine-tuned on labeled ECG-echocardiogram pairs for each SHD.

The third family is the publicly available EchoNext-Mini model, a single convolutional neural network developed at Columbia University that ingests ECG voltage-time waveforms and 7 auxiliary inputs (patient age, sex, ventricular rate, atrial rate, PR interval, QRS duration, and QTc) and produces probability outputs across 11 disease-specific nodes and a composite SHD node.^9,10^ The disease-specific nodes cover reduced left ventricular ejection fraction, increased left ventricular wall thickness, several moderate-to-severe valvular abnormalities (aortic, mitral, tricuspid, and pulmonary), right ventricular systolic dysfunction, pericardial effusion, elevated pulmonary artery pressure, and elevated tricuspid regurgitation velocity.

ECG signal and image preprocessing, and model inference strategy, are described in detail in the online supplement (**Supplementary Methods**).

### Test-Retest Reliability Assessment

Across all 3 health systems, we identified pairs of 12-lead ECGs recorded from the same individual between 1 and 30 days apart. Paired ECGs taken on the same day were excluded to avoid evaluating ECGs that were repeated due to acquisition errors. For individuals with more than one eligible ECG pair, one pair was randomly selected for inclusion. In YNHHS, we excluded any ECGs that were used in the model development cohort.

Test-retest reliability of continuous AI-ECG probability outputs was assessed using the concordance correlation coefficient (CCC), which captures both precision (correlation) and accuracy (deviation from the line of identity), with values closer to 1 indicating perfect concordance.^36^ Agreement of categorical screening results was assessed using percentage of ECG pairs with concordant screen status and Cohen’s kappa statistic, with screen-positive classification defined at the probability threshold for 90% sensitivity for each model during internal validation.

For the image-based ensemble composite, we additionally evaluated test-retest reliability across real-world ECG image acquisition modalities (EHR screenshots, smartphone photographs of monitor displays, and smartphone photographs of printed tracings) and across novel ECG plotting formats not encountered during training (variations in grid color, trace color, and lead arrangement, applied individually and in combination).

### Factors Associated with Model Discordance

To identify factors associated with discordant AI-ECG outputs, we developed multivariable logistic regression models at YNHHS with discordant screen status as the dependent variable. Discordance was defined as a change in binary screen classification between paired ECGs for the ensemble PRESENT-SHD model. Covariates included demographic factors (age, sex, race, and ethnicity), encounter type at ECG acquisition, time interval between paired ECGs, and ECG machine manufacturers.

### Predictive Significance of Discordant AI-ECG Results

To evaluate the predictive significance of discordant AI-ECG results, we identified cohorts of individuals without prevalent HF at baseline. A 1-year blanking period from the first recorded encounter in the electronic health record was applied to ascertain prevalent disease, and individuals with any HF diagnosis before or during this period were excluded. Following the blanking period at each site, individuals with paired ECGs recorded 30 to 90 days apart were included in this predictive analysis cohort. This time window balanced clinical proximity with temporal separation, minimizing immediate repeat ECGs while preserving a timeframe relevant to serial AI-ECG screening and follow-up.

Among individuals at risk, serial AI-ECG screen results were categorized by the screen classifications of paired ECGs into four groups: concordant negative, positive-to-negative change, negative-to-positive change, and concordant positive. The study outcome was defined as new-onset HF, defined as the first HF diagnosis recorded in the EHR. Follow-up began at the date of the second ECG in each pair and continued until the first HF diagnosis, death, or end of follow-up.

### Statistical Analysis

Continuous variables are reported as median (interquartile range, IQR) and categorical variables as counts (percentages). Test-retest reliability of continuous AI-ECG probability outputs was quantified using the CCC, accompanied by 95% confidence intervals (CIs) derived from 500 bootstrap iterations; agreement of categorical screen status was quantified using concordance percentages and Cohen’s kappa. Factors associated with discordant screen status were evaluated using multivariable logistic regression, with results expressed as odds ratios (ORs) with 95% CIs. The association of serial AI-ECG screen status with new-onset HF was evaluated using Cox proportional hazards models, with concordant negative screens as the reference category and sequential adjustment for demographics (age, sex, race, and ethnicity) and comorbidities (hypertension, diabetes mellitus, and obesity, each defined by the presence of any relevant diagnosis code prior to the second paired ECG); the proportional hazards assumption was evaluated by inspection of Schoenfeld residuals. Hazard ratios (HRs) are reported with 95% CIs.

All statistical analyses were performed using Python (version 3.11). Statistical significance was defined as a two-sided p-value less than 0.05. The study follows the Transparent Reporting of a Multivariable Prediction Model for Individual Prognosis or Diagnosis with Artificial Intelligence (TRIPOD+AI) guideline (**Supplementary Table 1**).

**Figure 1.**
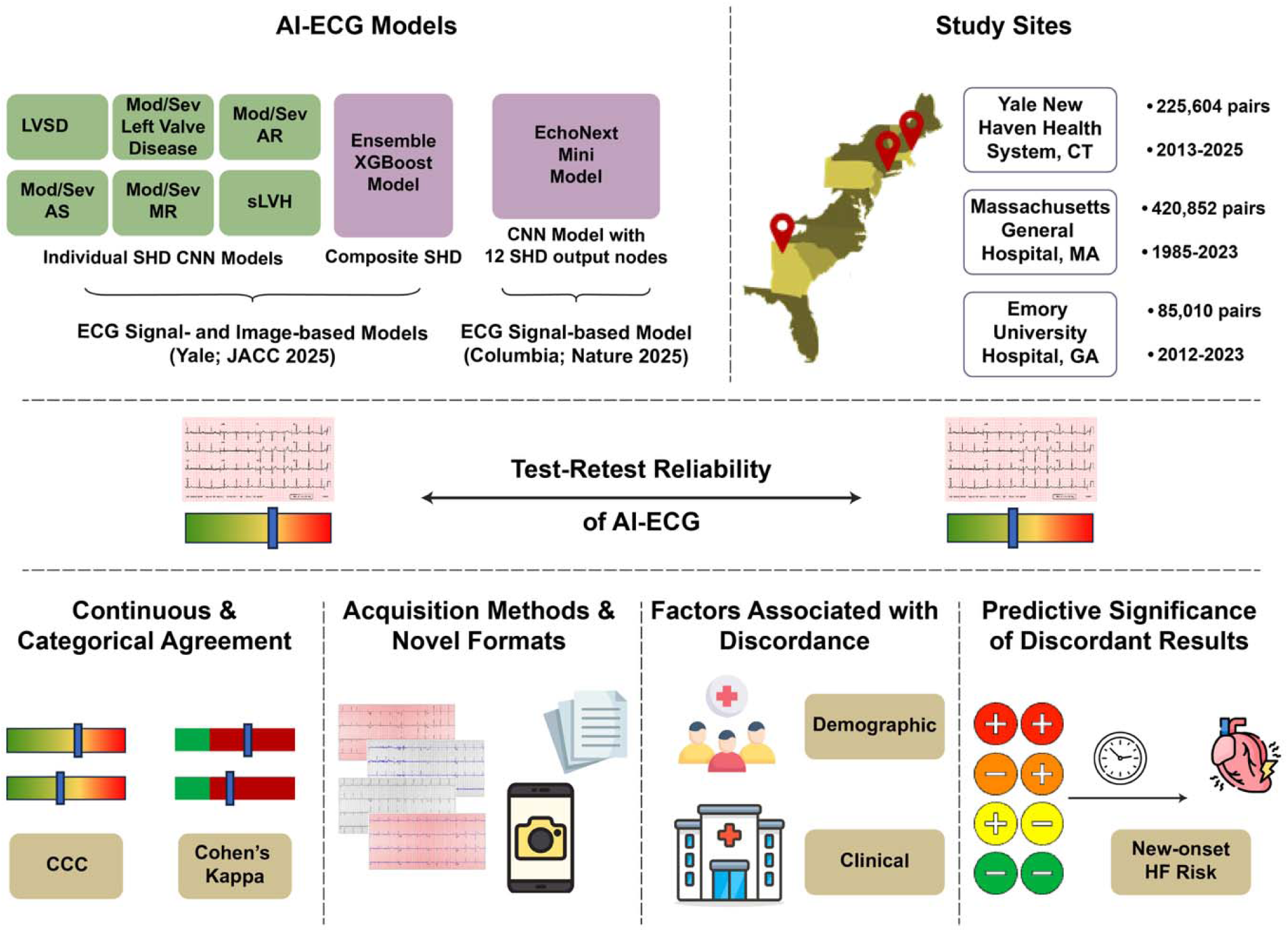
Study Design and Analytic Cohorts. Abbreviations: AR, aortic regurgitation; AS, aortic stenosis; CNN, convolutional neural network; ECG, electrocardiogram; FC, fully-connected layers; LVSD, left ventricular systolic dysfunction; MR, mitral regurgitation; SHD, structural heart diseases; sLVH, severe left ventricular hypertrophy; XGBoost, extreme gradient boosting.

## RESULTS

### Study Population

We included 225,604 ECG pairs at YNHHS, 420,852 at MGH, and 85,010 at EUH, with a median 5-6 days between paired ECGs at each site (**Supplementary Table 2**). Across sites, the median age was 65 years and women contributed 46-49% of ECG pairs. Racial and ethnic distributions differed substantially across sites: the proportion of patients identifying as non-Hispanic White ranged from 53% (EUH) to 67% (YNHHS), and the proportion identifying as non-Hispanic Black ranged from 6% (MGH) to 31% (EUH).

### Test-Retest Reliability: Continuous Agreement

Across disease-specific component models, CCCs spanned 0.77-0.85 for the signal-based Yale SHD models, 0.79-0.86 for the corresponding image-based models, and 0.50-0.97 for the disease-specific output nodes of the EchoNext-Mini model. The ensemble composite of each model family was uniformly more concordant than its components, with CCCs of 0.90 (95% CI, 0.89-0.90) for the signal-based ensemble, 0.90 (95% CI, 0.90-0.90) for the image-based ensemble, and 0.81 (95% CI, 0.81-0.81) for the EchoNext-Mini composite. The image- and signal-based model families yielded effectively identical CCCs across various disease-specific models, indicating that input modality did not materially alter reliability when development cohort and training strategy were held constant (**Figure 2; Supplementary Table 3**).

**Figure 2.**
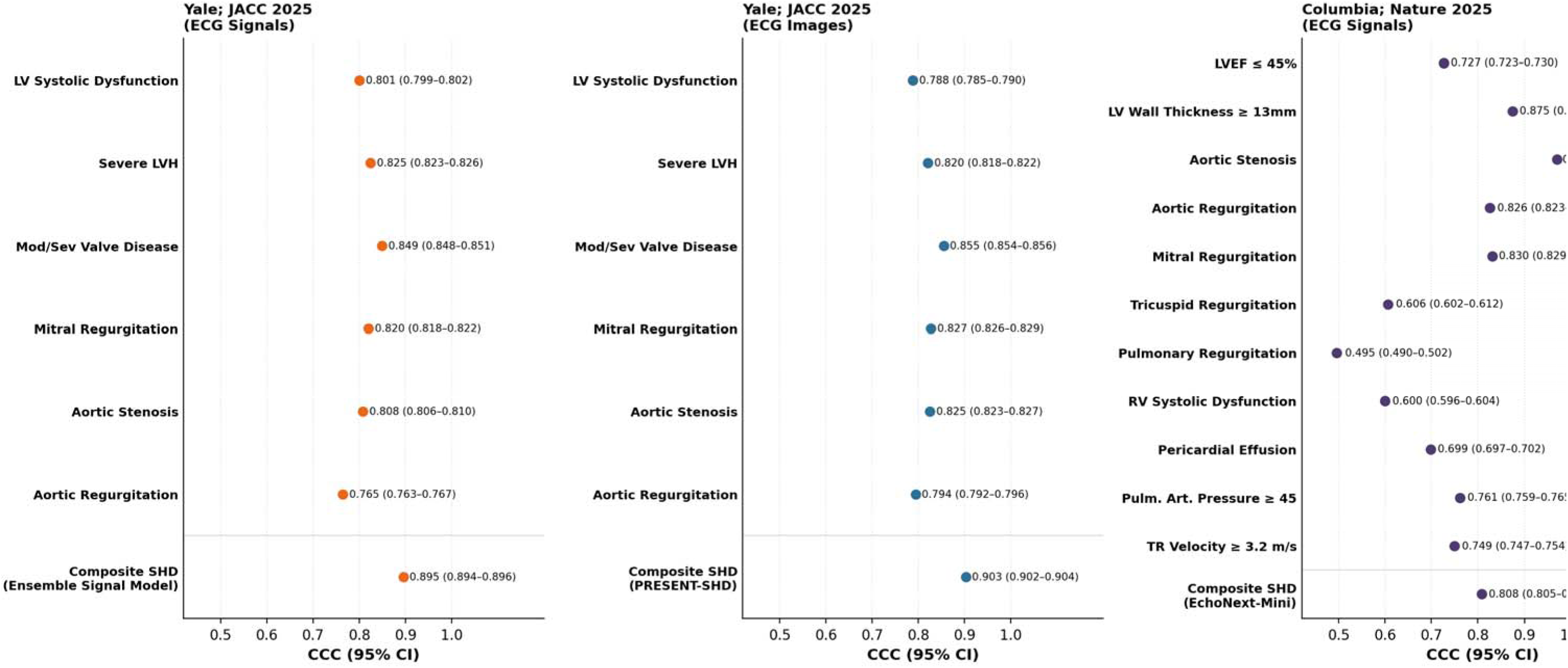
Test-Retest Concordance of Continuous AI-ECG Outputs across ECG Signal-Based and Image-Based Yale Structural Heart Disease, and EchoNext-Mini Model Families. Abbreviations: CNN, convolutional neural network; ECG, electrocardiogram; LVH, left ventricular hypertrophy; PRESENT-SHD, Practical Screening using Ensemble Machine Learning Strategy for Structural Heart Disease; XGBoost, extreme gradient boosting.

In external validation of the image-based models at MGH and EUH, the image-based composite preserved a high level of concordance, with CCCs of 0.88 (95% CI, 0.88-0.88) at MGH and 0.87 (0.86-0.87) at EUH; corresponding CCCs for disease-specific models ranged 0.76-0.82 at MGH and 0.76-0.83 at EUH (**Figure 2**).

### Test-Retest Reliability: Categorical Agreement

Across all 3 sites, the ensemble composite of the image-based SHD model reached higher categorical agreement between paired ECGs than its disease-specific components (**Supplementary Table 4**). Concordance of screen status ranged from 80-84% across disease-specific models at YNHHS, MGH, and EUH, compared with 89%, 87%, and 87% for the composite model (**Figure 3; Supplementary Table 4**). Cohen’s kappa followed a parallel pattern, ranging from approximately 0.56-0.68 for disease-specific models and from 0.72-0.78 for the composite across sites (**Supplementary Figure 1**).

**Figure 3.**
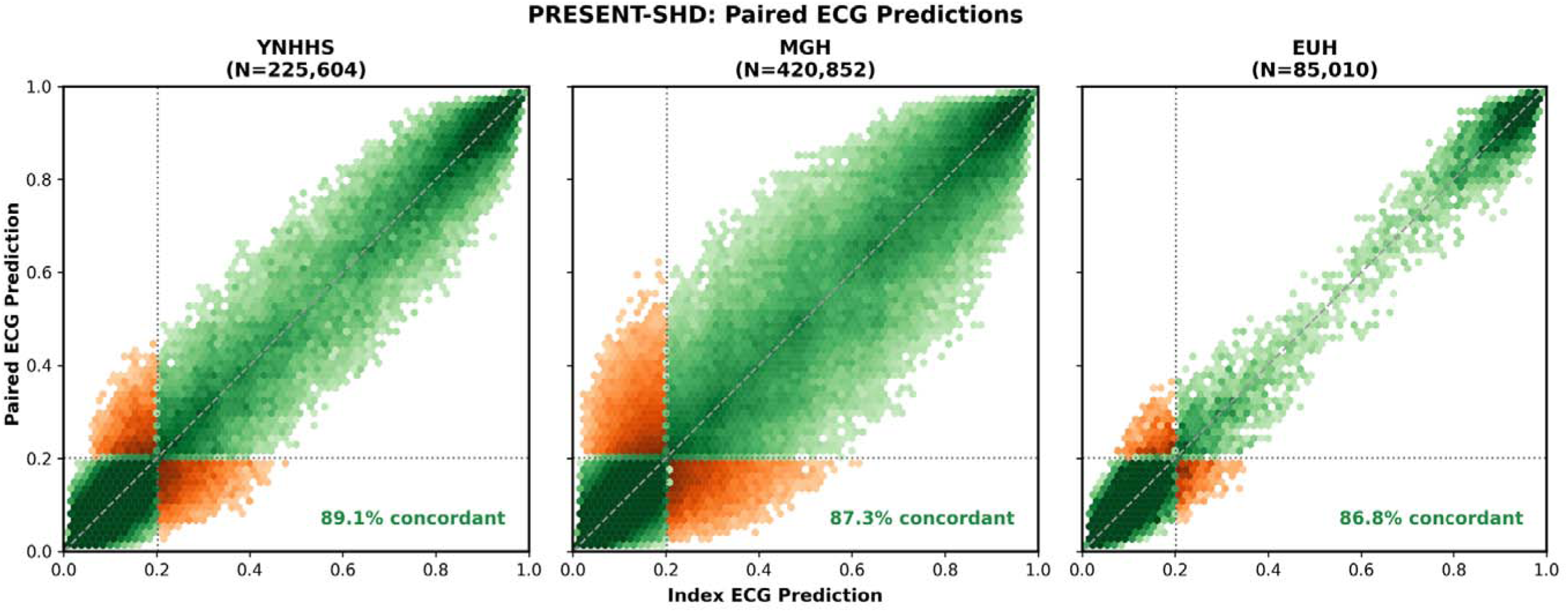
Paired-electrocardiogram probability scatter plots and concordance percentages for the ensemble image-based structural heart disease model across three health systems. Abbreviations: ECG, electrocardiogram; PRESENT-SHD, Practical Screening using Ensemble Machine Learning Strategy for Structural Heart Disease.

### Factors Associated with Discordant AI-ECG Outputs

In multivariable logistic regression, younger age was the strongest correlate of screen-status change between paired ECGs (**Figure 4**): patients younger than 65 years had 69% higher odds of discordance than those 65 years or older (OR 1.69 [95% CI, 1.65-1.74]). Sex and race or ethnicity showed modest associations with AI-ECG screen discordance in pairs (men versus women, OR 1.16 [95% CI, 1.13-1.19]; ORs ranging from 0.82 to 1.06 across racial and ethnic groups, compared with non-Hispanic White patients).

**Figure 4.**
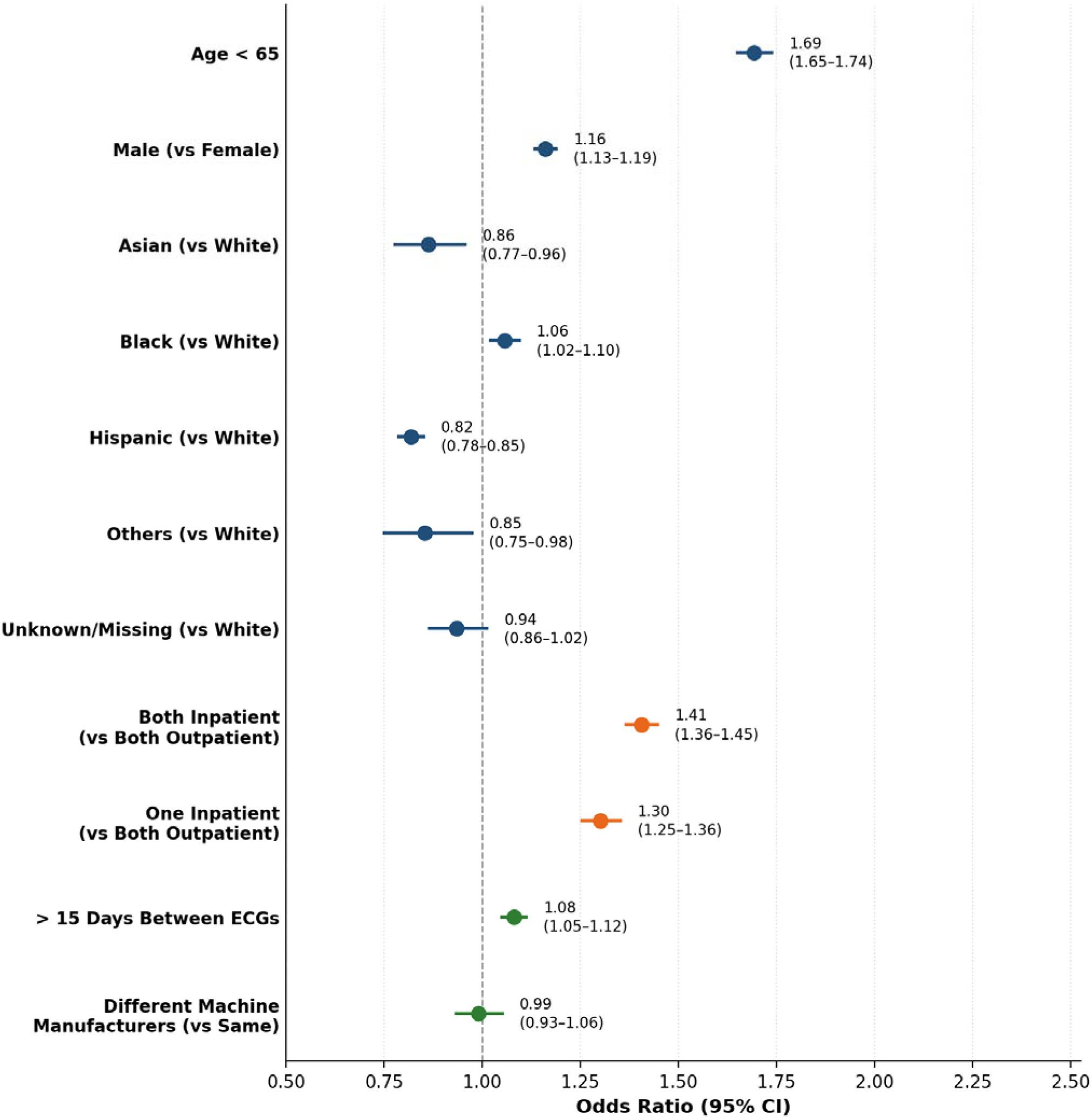
Patient- and encounter-level factors associated with discordant screen status in paired electrocardiograms. Abbreviations: CI, confidence interval; ECG, electrocardiogram; OR, odds ratio.

Adjusted for demographic and clinical characteristics, encounter setting was independently associated with discordance: compared with fully outpatient pairs, odds of discordance were higher when one ECG was acquired during an inpatient encounter (OR 1.30 [95% CI, 1.25-1.36]) and when both ECGs were acquired during inpatient encounters (OR 1.41 [95% CI, 1.36-1.45]; **Figure 4**). Pairs separated by more than 15 days had modestly higher odds than those within 15 days (OR 1.08 [95% CI, 1.05-1.12]), while pairs acquired with different ECG machine manufacturers did not differ from same-manufacturer pairs (OR 0.99 [95% CI, 0.93-1.06]).

### Reliability Across Real-World Acquisition Methods and ECG Image Layouts

The image-based composite SHD model preserved high test-retest reliability across real-world ECG acquisition modalities (EHR screenshots and smartphone photographs of monitor displays and printed tracings; CCC 0.96-0.97, concordance 89-92%) and across novel ECG plotting formats not encountered during training, including variations in grid color, trace color, and lead arrangement (CCC 0.96-0.998, concordance 93-99%; **Supplementary Table 5**). Bland-Altman analyses indicated minimal systematic bias and narrow limits of agreement across all evaluated formats (**Supplementary Figure 2**).

### Predictive Significance of Discordant AI-ECG Screens

In the cohort of individuals without prevalent HF at baseline who were screened with the ECG image-based ensemble composite, 39,517 at YNHHS, 22,940 at MGH, and 3,381 at EUH had paired ECGs eligible for the predictive analysis (**Supplementary Table 6**). Median follow-up was 4.7 years (IQR, 2.1-7.5) at YNHHS, 3.2 years (IQR, 1.5-5.1) at MGH, and 5.4 years (IQR, 1.8-6.4) at EUH, during which 1,876 (4.7%), 2,099 (9.1%), and 1,122 (33.2%) individuals developed new-onset HF, respectively. Pairs were categorized by serial screen status as concordant negative, positive-to-negative change, or negative-to-positive change and concordant positive statuses; the corresponding proportions were 44.5%, 10.7%, 5.5%, and 39.3%, at YNHHS; 50.0%, 7.3%, 6.9%, and 35.8% at MGH; and 32.4%, 7.7%, 7.7%, and 52.1% at EUH.

Relative to individuals with concordant negative screens, the serial-result categories showed a graded, increasing hazard for new-onset HF across all 3 sites in age-, sex-, and risk-factor-adjusted Cox models. Notably, positive-to-negative screen status changes carried a significantly elevated risk (HRs ranging 1.30-2.04 across sites) compared with concordant negative screens, followed by negative-to-positive changes (HRs 1.67-2.37) with greater risk, and concordant positive screens with the highest risk at each site (HRs 3.18-4.64; **Figure 5**). These associations were directionally consistent across sites, including after sequential adjustment for demographic and clinical covariates (**Supplementary Table 7**).

**Figure 5.**
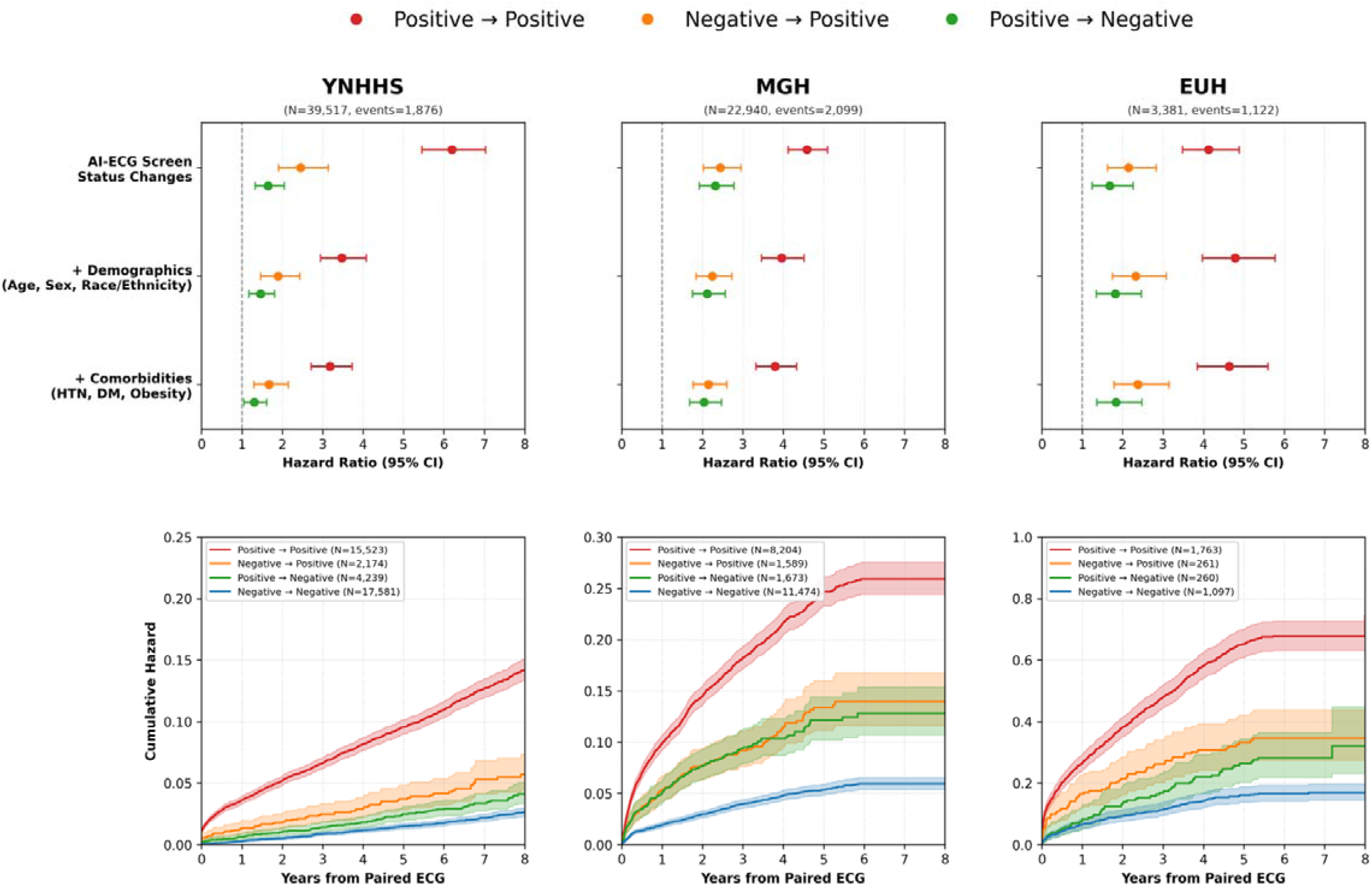
Risk of new-onset heart failure by serial change in screen status of artificial intelligence-enhanced electrocardiography across three health systems. (Top) Hazard ratios for each serial screen-status category from sequentially adjusted Cox proportional hazards models. (Bottom) Cumulative hazard curves from the fully adjusted Cox proportional hazards model. Abbreviations: CI, confidence interval; DM, type-2 diabetes mellitus; ECG, electrocardiogram; HR, hazard ratio; HTN, hypertension.

## DISCUSSION

In this multi-center evaluation of more than 730,000 paired ECGs across 3 geographically distinct US health systems, contemporary AI-ECG models for SHD detection had high test-retest reliability when repeated under similar clinical conditions. Ensemble composites that aggregated multiple disease-specific outputs yielded more stable results between paired ECGs than the underlying disease-specific component models, a pattern that was reproduced across signal-based and image-based models, and the single-network multi-output composite implementation. Reliability of the image-based models was maintained across real-world image acquisition strategies (screenshots, photographs, printouts) and novel ECG plotting formats. Among patient and encounter characteristics, younger age and inpatient ECG acquisition were the principal correlates of a change in screen status between paired ECGs; differences by sex, race or ethnicity, ECG machine manufacturer, and time between ECGs were modest. Serial screen-status patterns carried an independent predictive signal, such that among individuals without prevalent HF, new-onset HF risk followed a graded pattern from concordant negative through concordant positive screens, with positive-to-negative and negative-to-positive changes carrying intermediate hazards.

The adoption of AI-ECG tools into clinical workflows has accelerated alongside an expanding FDA regulatory framework for AI-enabled software-based clinical decision support systems.^37,38^ While over 1,000 AI-based medical device authorizations have been issued to date, post-market surveillance data indicate that a meaningful proportion of authorized AI devices have been recalled or corrected.^39,40^ Properties beyond initial validation accuracy, such as test-retest consistency, require systematic characterization before widespread deployment. The scientific AI-ECG literature has largely focused on discrimination and calibration in cross-sectional cohorts, using one ECG per individual.^2,5,7–9,24–28^ This approach is essential for model validation, but it does not fully capture how these tools behave when applied repeatedly to the same patient. A test that performs well at the population level but varies substantially on repeat testing can create uncertainty for clinicians and patients. A useful parallel is clinical laboratory medicine, where the coefficient of variation is reported alongside sensitivity and specificity. This is particularly important for AI-ECG tools, which are increasingly being considered for screening, triage, and longitudinal risk assessment. Excessive variation can prompt unnecessary follow-up, whereas excessive stability can create false reassurance in the setting of evolving disease. Our findings provide an empirical assessment of where contemporary AI-ECG models fall on this stability spectrum, across 3 health systems and 3 distinct model families, positioning test-retest reliability as a complementary, deployment-relevant benchmark alongside discrimination and calibration.

Ensemble composites were more reliable than disease-specific component models across all 3 AI-ECG families. Aggregating multiple component outputs can average idiosyncratic responses to acquisition-level variation affecting any single disease-specific model, thereby stabilizing the composite prediction. This pattern has direct clinical implications. Because a composite SHD model targets a broader group of clinically important conditions sharing a common confirmatory test, a transthoracic echocardiography, combining multiple disease-specific signals can improve both yield and reliability of SHD screening. The near-identical reliability of signal- and image-based models is notable given that these architectures process different representations of the same underlying electrical activity.^5,41^ This modality equivalence has practical implications for large-scale deployment. Health systems implementing AI-ECG screening across mixed acquisition environments can expect comparable test-retest stability across modalities. The consistency of outputs across image layouts and acquisition methods further supports their use in varied deployment contexts.

Among patient and encounter characteristics, 2 factors showed substantially elevated odds of a change in screen status between paired ECGs: younger age and inpatient acquisition context. The association with younger age may reflect clinical selection among younger individuals who undergo ECG testing. These individuals may have ECGs obtained for clinical symptoms, pre-procedural evaluation, or acute concerns. The association with inpatient ECG acquisition reflects expected physiologic variability in hospitalized patients due to short-term changes in volume status, medications, rhythm, ischemia, or acute illness that alter ECG features over short intervals. Acute clinical context therefore modifies interpretation of serial AI-ECG results and warrants context-aware deployment strategies. Discordance was not higher for pairs acquired on ECG machines from different manufacturers than for same-manufacturer pairs, supporting multi-site deployment without requiring uniform ECG hardware.

Among cardiac biomarkers, natriuretic peptides and high-sensitivity troponins are routinely interpreted in the context of prior values, with the direction and magnitude of change informing clinical decision-making alongside absolute level. Our findings demonstrate that serial AI-ECG, capturing a broad phenotypic signature, can similarly provide longitudinal information, extending the clinical utility of AI-ECG beyond a single-time-point binary classification.^14,35^ Consistent across all 3 sites, the graded association between serial screen-status trajectory and HF risk suggests that these changes reflect evolving cardiovascular biology rather than random output fluctuation, especially when applied using reliable composite models. In practice, a new positive AI-ECG screen after a prior negative result may warrant clinical attention, confirmatory imaging, or closer preventive monitoring, rather than being treated as a technical artifact. We therefore propose that test-retest reliability be reported alongside discrimination and calibration as a standard implementation benchmark in AI-ECG validation studies. This would enable a more complete characterization of model behavior as these tools scale from academic development into broad clinical deployment.

This study has limitations that merit consideration. First, the analytic cohorts comprised individuals who underwent 2 ECGs within 30 days, which selects toward patients with greater healthcare utilization. However, the consistency of findings across 3 health systems with distinct patient populations and clinical practices makes site-specific selection less likely to drive the observed patterns. Second, although a 1-day minimum interval reduced inclusion of immediate technical repeats, some pairs may have included repeats motivated by artifact. This would bias estimates toward apparent discordance. Thus, the high observed reliability is likely a conservative estimate of true reliability. Third, follow-up duration and HF event rates varied across sites in the predictive analyses, which may reflect differences in referral patterns and EHR capture. Nonetheless, the directional consistency of the predictive associations across sites despite this heterogeneity supports robustness of the findings. Finally, although models were evaluated across format variations and real-world image acquisition modalities, prospective deployment studies will be needed to characterize AI-ECG behavior in active screening programs and in settings beyond the academic health systems studied here.

## CONCLUSION

Across diverse health systems, AI-ECG models for SHD had high test-retest reliability, with composite ensemble models showing the greatest stability. Discordance in AI-ECG results between serial ECGs was associated with an elevated risk of new-onset HF, supporting the reliability and longitudinal screen-status changes as clinically informative implementation benchmarks for AI-ECG.

## Data Availability

Individual-level data from the Yale-New Haven Health System cannot be shared publicly due to HIPAA regulations enforced by the Yale Institutional Review Board. The image-based AI-ECG models developed at Yale are publicly accessible for research use on the Cardiovascular Data Science (CarDS) Lab website, and the EchoNext-Mini model is available at https://github.com/PierreElias/IntroECG. Study code will be publicly released on GitHub upon publication.

## ABBREVIATIONS LIST

AI-ECG: artificial intelligence-enhanced electrocardiography
CCC: concordance correlation coefficient
CI: confidence intervals
CNN: convolutional neural network
EHR: electronic health record
EUH: Emory University Hospital
HF: heart failure
HR: hazard ratio
IQR: interquartile range
LVSD: left ventricular systolic dysfunction
MGH: Massachusetts General Hospital
OR: odds ratio
PRESENT-SHD: Practical scREening using ENsemble machine learning sTrategy for Structural Heart Disease
XGBoost: extreme gradient boosting
YNHHS: Yale-New Haven Health System

