## Supplementary material for "Test-Retest Reliability of Artificial Intelligence-Enhanced Electrocardiography: A Multi-Center Study": Online Supplement

### Supplement Materials

### SUPPLEMENTARY METHODS

#### ECG Image Generation and Format Experiments

For image-based AI-ECG models, ECG waveforms were plotted as images in a standard clinical format with a voltage calibration of 10 mm/mV. Limb and precordial leads were arranged in four columns of 2.5 seconds each, representing leads I, II, and III; aVR, aVL, and aVF; V1, V2, and V3; and V4, V5, and V6. A 10-second recording of the lead I signal was included as a rhythm strip. All images were converted to grayscale and downsampled to 300 × 300 pixels for model inference.[1–3]

To evaluate model reliability across real-world ECG image acquisition modalities, we tested three acquisition methods: screenshots of ECGs exported from the electronic health record (EHR), smartphone photographs of ECG images displayed on computer monitors, and smartphone photographs of ECGs printed on standard paper. We additionally tested models across novel ECG plotting formats not encountered during training, including variations in background grid color, ECG trace color, and lead arrangement, applied individually and in combination.[2]

#### EchoNext-Mini Model Inference

The EchoNext-Mini model was deployed in its publicly available form.[4,5] The model ingests 12-lead ECG voltage-time waveforms along with patient age, sex, and standard ECG-derived intervals (ventricular rate, atrial rate, PR interval, QRS duration, and QT/QTc) as auxiliary inputs. The model produces continuous probability outputs across 11 disease-specific nodes, covering reduced left ventricular ejection fraction (≤45%), increased left ventricular wall thickness (≥13 mm), moderate-to-severe valvular abnormalities (aortic stenosis, aortic regurgitation, mitral regurgitation, tricuspid regurgitation, and pulmonary regurgitation), right ventricular systolic dysfunction, pericardial effusion, pulmonary artery pressure ≥45 mmHg, and tricuspid regurgitation velocity ≥3.2 m/s, and a composite SHD output node. We applied EchoNext-Mini to the YNHHS ECG-pair cohort without additional fine-tuning.

### SUPPLEMENTARY FIGURES

**Supplementary Figure 1. Cohen's kappa for test-retest agreement of categorical screen status across the image-based Artificial Intelligence-enhanced Electrocardiography models at three health systems.** Abbreviations: CI, confidence intervals; CNN, convolutional neural network; EUH, Emory University Hospital; LV, left ventricular; LVH, left ventricular hypertrophy; LVSD, left ventricular systolic dysfunction; MGH, Massachusetts General Hospital; Mod/Sev, moderate to severe; PRESENT-SHD, Practical Screening using Ensemble Machine Learning Strategy for Structural Heart Disease; SHD, structural heart disease; YNHHS, Yale-New Haven Health System.

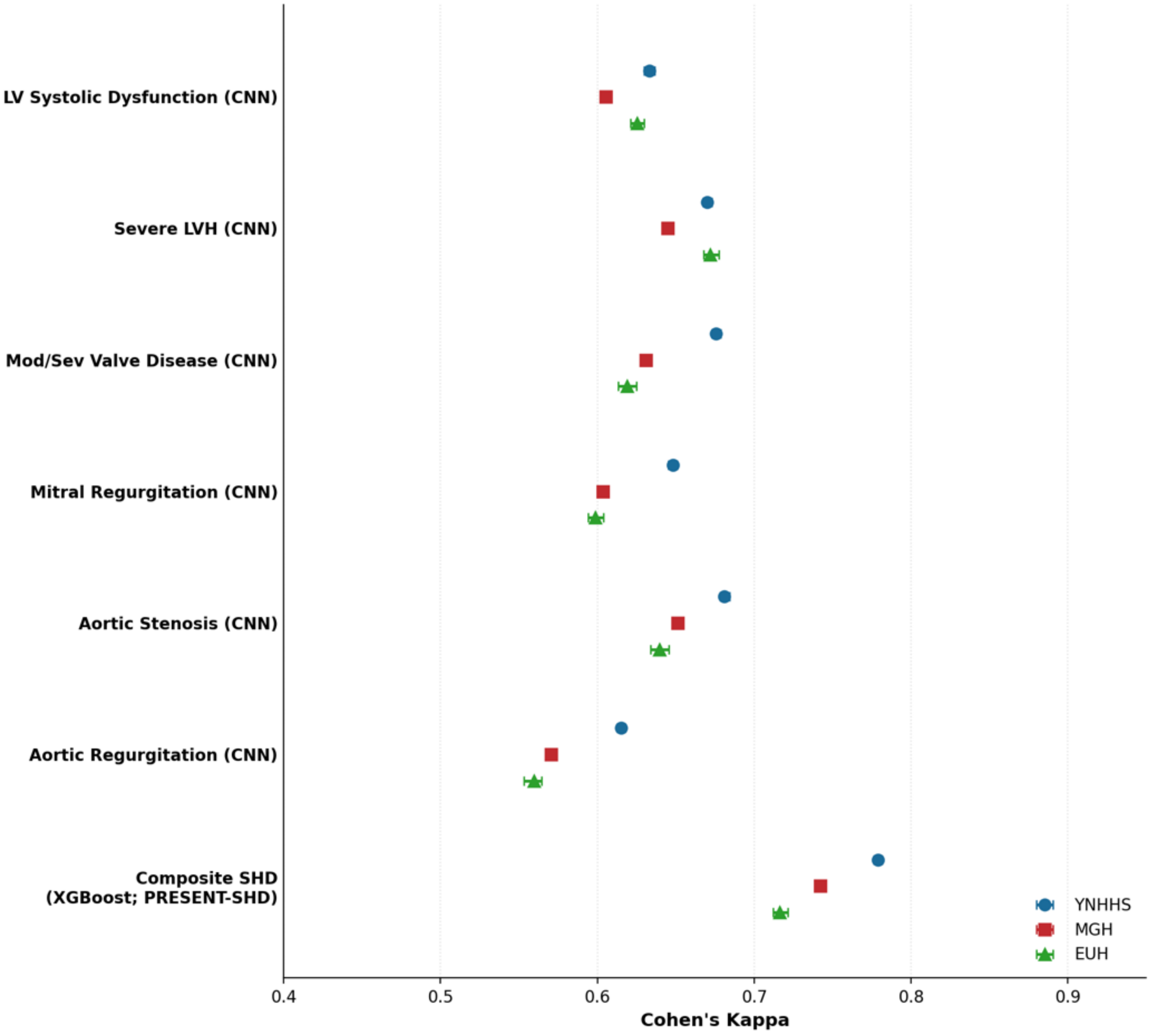

**Supplementary Figure 2. Bland-Altman analyses comparing paired-ECG probability outputs of the image-based ensemble composite (PRESENT-SHD) across (A) real-world image acquisition modalities and (B) novel ECG plotting formats, relative to standard plotted images.** Abbreviations: ECG, electrocardiogram; EHR, electronic health record; PRESENT-SHD, Practical Screening using Ensemble Machine Learning Strategy for Structural Heart Disease.

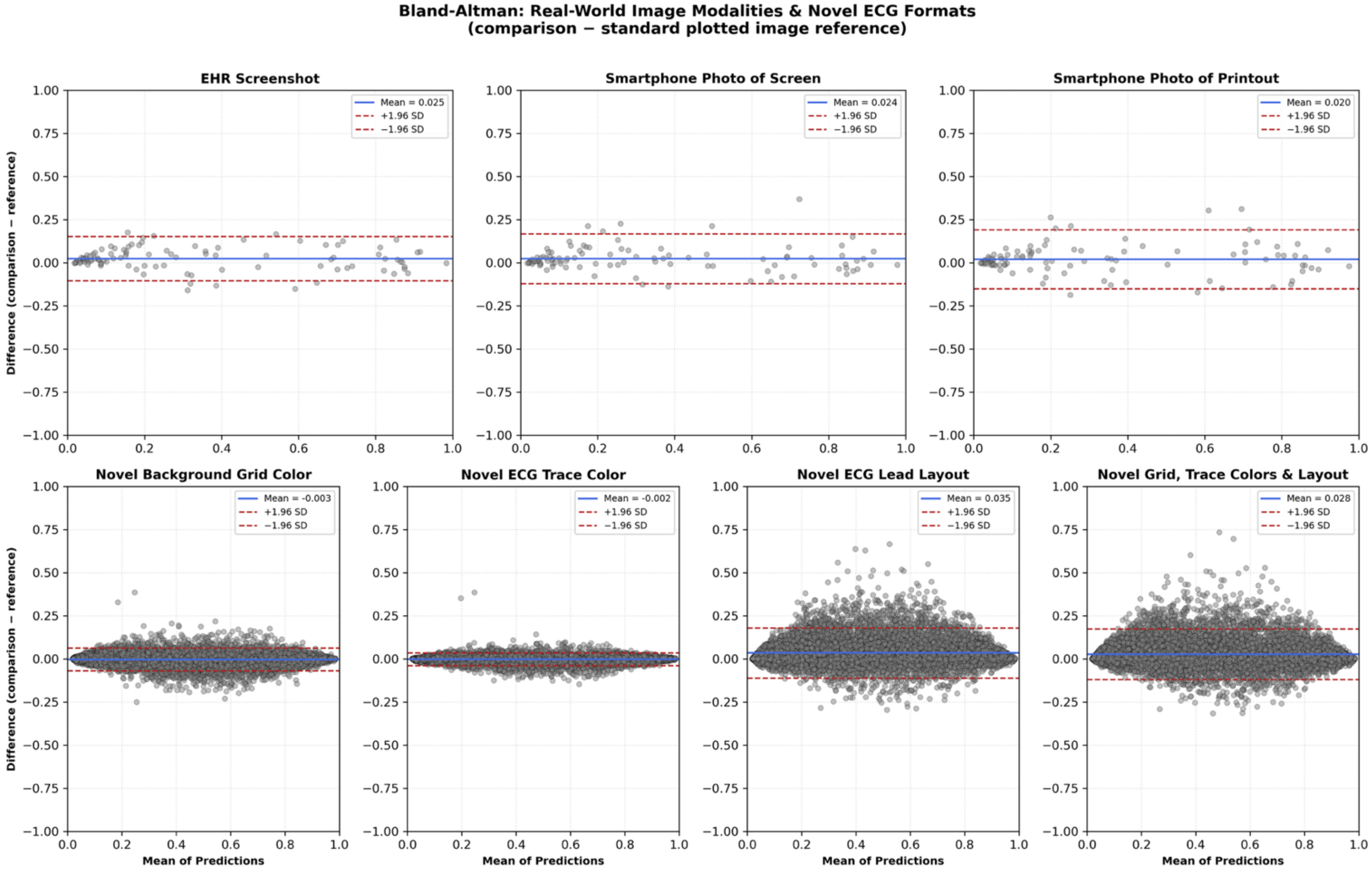

### Supplementary Table 1. TRIPOD-AI Checklist.

TRIPOD+AI

Version: 11-January-2024

| Section/Topic | Item | Development / evaluation[1] | Checklist item | Reported on page |
|---|---|---|---|---|
| **TITLE** | | | | |
| *Title* | 1 | D;E | Identify the study as developing or evaluating the performance of a multivariable prediction model, the target population, and the outcome to be predicted | 1 |
| **ABSTRACT** | | | | |
| *Abstract* | 2 | D;E | See TRIPOD+AI for Abstracts checklist | 3-4 |
| **INTRODUCTION** | | | | |
| *Background* | 3a | D;E | Explain the healthcare context (including whether diagnostic or prognostic) and rationale for developing or evaluating the prediction model, including references to existing models | 7-8 |
| | 3b | D;E | Describe the target population and the intended purpose of the prediction model in the context of the care pathway, including its intended users (e.g., healthcare professionals, patients, public) | 7-8 |
| | 3c | D;E | Describe any known health inequalities between sociodemographic groups | N/A |
| *Objectives* | 4 | D;E | Specify the study objectives, including whether the study describes the development or validation of a prediction model (or both) | 8 |
| **METHODS** | | | | |
| *Data* | 5a | D;E | Describe the sources of data separately for the development and evaluation datasets (e.g., randomised trial, cohort, routine care or registry data), the rationale for using these data, and representativeness of the data | 8-9 |
| | 5b | D;E | Specify the dates of the collected participant data, including start and end of participant accrual; and, if applicable, end of follow-up | 8-9 |
| *Participants* | 6a | D;E | Specify key elements of the study setting (e.g., primary care, secondary care, general population) including the number and location of centres | 8-9 |
| | 6b | D;E | Describe the eligibility criteria for study participants | 11-12 |
| | 6c | D;E | Give details of any treatments received, and how they were handled during model development or evaluation, if relevant | N/A |
| *Data preparation* | 7 | D;E | Describe any data pre-processing and quality checking, including whether this was similar across relevant sociodemographic groups | 9-10; Supp |
| *Outcome* | 8a | D;E | Clearly define the outcome that is being predicted and the time horizon, including how and when assessed, the rationale for choosing this outcome, and whether the method of outcome assessment is consistent across sociodemographic groups | 11-13 |
| | 8b | D;E | If outcome assessment requires subjective interpretation, describe the qualifications and demographic characteristics of the outcome assessors | N/A |
| | 8c | D;E | Report any actions to blind assessment of the outcome to be predicted | N/A |
| *Predictors* | 9a | D | Describe the choice of initial predictors (e.g., literature, previous models, all available predictors) and any pre-selection of predictors before model building | N/A |
| | 9b | D;E | Clearly define all predictors, including how and when they were measured (and any actions to blind assessment of predictors for the outcome and other predictors) | 11-13 |
| | 9c | D;E | If predictor measurement requires subjective interpretation, describe the qualifications and demographic characteristics of the predictor assessors | N/A |
| *Sample size* | 10 | D;E | Explain how the study size was arrived at (separately for development and evaluation), and justify that the study size was sufficient to answer the research question. Include details of any sample size calculation | 10; 13 |
| *Missing data* | 11 | D;E | Describe how missing data were handled. Provide reasons for omitting any data | Supp |
| *Analytical methods* | 12a | D | Describe how the data were used (e.g., for development and evaluation of model performance) in the analysis, including whether the data were partitioned, considering any sample size requirements | N/A |
| | 12b | D | Depending on the type of model, describe how predictors were handled in the analyses (functional form, rescaling, transformation, or any standardisation). | N/A |
| | 12c | D | Specify the type of model, rationale[2], all model-building steps, including any hyperparameter tuning, and method for internal validation | N/A |
| | 12d | D;E | Describe if and how any heterogeneity in estimates of model parameter values and model performance was handled and quantified across clusters (e.g., hospitals, countries). See TRIPOD-Cluster for additional considerations[3] | 11-13 |
| | 12e | D;E | Specify all measures and plots used (and their rationale) to evaluate model performance (e.g., discrimination, calibration, clinical utility) and, if relevant, to compare multiple models | 11-13 |
| | 12f | E | Describe any model updating (e.g., recalibration) arising from the model evaluation, either overall or for particular sociodemographic groups or settings | N/A |
| | 12g | E | For model evaluation, describe how the model predictions were calculated (e.g., formula, code, object, application programming interface) | 11-13 |
| *Class imbalance* | 13 | D;E | If class imbalance methods were used, state why and how this was done, and any subsequent methods to recalibrate the model or the model predictions | N/A |
| *Fairness* | 14 | D;E | Describe any approaches that were used to address model fairness and their rationale | 13 |
| *Model output* | 15 | D | Specify the output of the prediction model (e.g., probabilities, classification). Provide details and rationale for any classification and how the thresholds were identified | N/A |

[1] D=items relevant only to the development of a prediction model; E=items relating solely to the evaluation of a prediction model; D;E=items applicable to both the development and evaluation of a prediction model
[2] Separately for all model building approaches.
[3] TRIPOD-Cluster is a checklist of reporting recommendations for studies developing or validating models that explicitly account for clustering or explore heterogeneity in model performance (eg, at different hospitals or centres). Debray et al, BMJ 2023; 380: e071018 [DOI: 10.1136/bmj-2022-071018]

| | | | | |
|---|---|---|---|---|
| *Training versus evaluation* | 16 | D;E | Identify any differences between the development and evaluation data in healthcare setting, eligibility criteria, outcome, and predictors | N/A |
| *Ethical approval* | 17 | D;E | Name the institutional research board or ethics committee that approved the study and describe the participant-informed consent or the ethics committee waiver of informed consent | 8 |
| **OPEN SCIENCE** | | | | |
| *Funding* | 18a | D;E | Give the source of funding and the role of the funders for the present study | 2 |
| *Conflicts of interest* | 18b | D;E | Declare any conflicts of interest and financial disclosures for all authors | 2 |
| *Protocol* | 18c | D;E | Indicate where the study protocol can be accessed or state that a protocol was not prepared | Not Preprared |
| *Registration* | 18d | D;E | Provide registration information for the study, including register name and registration number, or state that the study was not registered | Not Preprared |
| *Data sharing* | 18e | D;E | Provide details of the availability of the study data | 2 |
| *Code sharing* | 18f | D;E | Provide details of the availability of the analytical code[4] | 2 |
| **PATIENT & PUBLIC INVOLVEMENT** | | | | |
| *Patient & Public Involvement* | 19 | D;E | Provide details of any patient and public involvement during the design, conduct, reporting, interpretation, or dissemination of the study or state no involvement. | N/A |
| **RESULTS** | | | | |
| *Participants* | 20a | D;E | Describe the flow of participants through the study, including the number of participants with and without the outcome and, if applicable, a summary of the follow-up time. A diagram may be helpful. | 13; Supp |
| | 20b | D;E | Report the characteristics overall and, where applicable, for each data source or setting, including the key dates, key predictors (including demographics), treatments received, sample size, number of outcome events, follow-up time, and amount of missing data. A table may be helpful. Report any differences across key demographic groups. | 13; Supp |
| | 20c | E | For model evaluation, show a comparison with the development data of the distribution of important predictors (demographics, predictors, and outcome). | Supp |
| *Model development* | 21 | D;E | Specify the number of participants and outcome events in each analysis (e.g., for model development, hyperparameter tuning, model evaluation) | 13; Supp |
| *Model specification* | 22 | D | Provide details of the full prediction model (e.g., formula, code, object, application programming interface) to allow predictions in new individuals and to enable third-party evaluation and implementation, including any restrictions to access or re-use (e.g., freely available, proprietary)[5] | N/A |
| *Model performance* | 23a | D;E | Report model performance estimates with confidence intervals, including for any key subgroups (e.g., sociodemographic). Consider plots to aid presentation. | Supp |
| | 23b | D;E | If examined, report results of any heterogeneity in model performance across clusters. See TRIPOD Cluster for additional details[3]. | 29; Supp |
| *Model updating* | 24 | E | Report the results from any model updating, including the updated model and subsequent performance | N/A |
| **DISCUSSION** | | | | |
| *Interpretation* | 25 | D;E | Give an overall interpretation of the main results, including issues of fairness in the context of the objectives and previous studies | 17-20 |
| *Limitations* | 26 | D;E | Discuss any limitations of the study (such as a non-representative sample, sample size, overfitting, missing data) and their effects on any biases, statistical uncertainty, and generalizability | 20 |
| *Usability of the model in the context of current care* | 27a | D | Describe how poor quality or unavailable input data (e.g., predictor values) should be assessed and handled when implementing the prediction model | N/A |
| | 27b | D | Specify whether users will be required to interact in the handling of the input data or use of the model, and what level of expertise is required of users | N/A |
| | 27c | D;E | Discuss any next steps for future research, with a specific view to applicability and generalizability of the model | 17-20 |

From: Collins GS, Moons KGM, Dhiman P, et al. *BMJ* 2024;385:e078378. doi:10.1136/bmj-2023-078378

[4] This relates to the analysis code, for example, any data cleaning, feature engineering, model building, evaluation.
[5] This relates to the code to implement the model to get estimates of risk for a new individual.

**Supplementary Table 2. Demographic and clinical characteristics for patients with paired electrocardiograms across study sites.** Abbreviations: ECG, electrocardiogram; DM, type-2 diabetes mellitus; HTN, hypertension.

| **Model** | **YNHHS** | **MGH** | **EUH** |
|---|---|---|---|
| **Number of Patients** | 225604 | 420852 | 85010 |
| **Age (years; Median [IQR])** | 65.0 [50.0,78.0] | 65.1 [52.3,76.3] | 65.0 [53.6,74.4] |
| **Female Sex (%)** | 110759 (49.1) | 196539 (46.7) | 39323 (46.3) |
| **Race/Ethnicity (%)** | | | |
| **Non-Hispanic White** | 151395 (67.1) | 279484 (66.4) | 45010 (52.9) |
| **Non-Hispanic Black** | 30350 (13.5) | 26391 (6.3) | 26496 (31.2) |
| **Hispanic** | 25470 (11.3) | 25135 (6.0) | 343 (0.4) |
| **Non-Hispanic Asian** | 3557 (1.6) | 9853 (2.3) | 1389 (1.6) |
| **Non-Hispanic Others** | 6987 (3.1) | 57573 (13.7) | 4020 (4.7) |
| **Unknown** | 7845 (3.5) | 20029 (4.8) | 1302 (1.5) |
| **HTN (%)** | 150308 (66.6) | 42642 (10.1) | 3103 (3.7) |
| **DM (%)** | 64578 (28.6) | 17408 (4.1) | 3120 (3.7) |
| **Obesity (%)** | 56599 (25.1) | 11852 (2.8) | 1353 (1.6) |
| **Delta Between ECGs (days; median [IQR])** | 5.0 [1.0,15.0] | 6.0 [2.0,14.0] | 5.0 [1.0,13.0] |
| **ECG Setting (%)** | - | - | - |
| **Both Outpatient** | 81,945 (36.3) | - | - |
| **Both Inpatient** | 108,544 (48.1) | - | - |
| **One Inpatient** | 35,115 (15.6) | - | - |

**Supplementary Table 3. Model architectures, inference inputs, and test-retest concordance correlation coefficients for artificial intelligence-enhanced electrocardiography models evaluated at the Yale-New Haven Health System.** Abbreviations: CCC, concordance correlation coefficient; CI, confidence intervals; CNN, convolutional neural network; ECG, electrocardiograms; LV, left ventricular; LVEF, left ventricular ejection fraction; LVH, left ventricular hypertrophy; Mod/Sev, moderate to severe; PRESENT-SHD, Practical Screening using Ensemble Machine Learning Strategy for Structural Heart Disease; Pulm. Art., pulmonary artery; RV, right ventricular; SHD, structural heart disease; TR, tricuspid regurgitation; XGBoost, extreme gradient boosting.

| Model Type | Disease Label | Model Type | Required ECG Inputs | CCC (95% CI) |
|---|---|---|---|---|
| **Yale SHD Signal Models (JACC 2025)** | LV Systolic Dysfunction | CNN | ECG waveform signal | 0.801 (0.799-0.802) |
| | Severe LVH | CNN | ECG waveform signal | 0.825 (0.823-0.826) |
| | Mod/Sev Valve Disease | CNN | ECG waveform signal | 0.849 (0.848-0.851) |
| | Mitral Regurgitation | CNN | ECG waveform signal | 0.820 (0.818-0.822) |
| | Aortic Stenosis | CNN | ECG waveform signal | 0.808 (0.806-0.810) |
| | Aortic Regurgitation | CNN | ECG waveform signal | 0.765 (0.763-0.767) |
| | Composite SHD | XGBoost | ECG waveform signal, Age, Sex | 0.895 (0.894-0.896) |
| **Yale SHD Image Models (JACC 2025)** | LV Systolic Dysfunction | CNN | ECG Image | 0.788 (0.785-0.790) |
| | Severe LVH | CNN | ECG Image | 0.820 (0.818-0.822) |
| | Mod/Sev Valve Disease | CNN | ECG Image | 0.855 (0.854-0.856) |
| | Mitral Regurgitation | CNN | ECG Image | 0.827 (0.826-0.829) |
| | Aortic Stenosis | CNN | ECG Image | 0.825 (0.823-0.827) |
| | Aortic Regurgitation | CNN | ECG Image | 0.794 (0.792-0.796) |
| | Composite SHD (PRESENT-SHD) | XGBoost | ECG Image, Age, Sex | 0.903 (0.902-0.904) |
| **Columbia SHD EchoNext-mini Model (Nature 2025)** | LVEF ≤ 45% | CNN output node | ECG waveform signal, Age, Sex, Ventricular Rate, Atrial Rate, PR interval, QRS Duration, QT/QTc | 0.727 (0.723-0.730) |
| | LV Wall Thickness ≥ 13mm | CNN output node | ECG waveform signal, Age, Sex, Ventricular Rate, Atrial Rate, PR interval, QRS Duration, QT/QTc | 0.875 (0.873-0.877) |
| | Aortic Stenosis | CNN output node | ECG waveform signal, Age, Sex, Ventricular Rate, Atrial Rate, PR interval, QRS Duration, QT/QTc | 0.970 (0.969-0.970) |
| | Aortic Regurgitation | CNN output node | ECG waveform signal, Age, Sex, Ventricular Rate, Atrial Rate, PR interval, QRS Duration, QT/QTc | 0.826 (0.823-0.829) |
| | Mitral Regurgitation | CNN output node | ECG waveform signal, Age, Sex, Ventricular Rate, Atrial Rate, PR interval, QRS Duration, QT/QTc | 0.830 (0.829-0.833) |
| | Tricuspid Regurgitation | CNN output node | ECG waveform signal, Age, Sex, Ventricular Rate, Atrial Rate, PR interval, QRS Duration, QT/QTc | 0.606 (0.602-0.612) |
| | Pulmonary Regurgitation | CNN output node | ECG waveform signal, Age, Sex, Ventricular Rate, Atrial Rate, PR interval, QRS Duration, QT/QTc | 0.495 (0.490-0.502) |
| | RV Systolic Dysfunction | CNN output node | ECG waveform signal, Age, Sex, Ventricular Rate, Atrial Rate, PR interval, QRS Duration, QT/QTc | 0.600 (0.596-0.604) |
| | Pericardial Effusion | CNN output node | ECG waveform signal, Age, Sex, Ventricular Rate, Atrial Rate, PR interval, QRS Duration, QT/QTc | 0.699 (0.697-0.702) |
| | Pulm. Art. Pressure ≥ 45 | CNN output node | ECG waveform signal, Age, Sex, Ventricular Rate, Atrial Rate, PR interval, QRS Duration, QT/QTc | 0.761 (0.759-0.765) |
| | TR Velocity ≥ 3.2 m/s | CNN output node | ECG waveform signal, Age, Sex, Ventricular Rate, Atrial Rate, PR interval, QRS Duration, QT/QTc | 0.749 (0.747-0.754) |
| | Composite SHD (EchoNext-Mini) | CNN output node | ECG waveform signal, Age, Sex, Ventricular Rate, Atrial Rate, PR interval, QRS Duration, QT/QTc | 0.808 (0.805-0.811) |

**Supplementary Table 4. Concordance percentages and Cohen's kappa for test-retest agreement of categorical screen status across image-based artificial intelligence-enhanced electrocardiography models and three health systems.** Abbreviations: CI, confidence intervals; CNN, convolutional neural network; ECG, electrocardiograms; EUH, Emory University Hospital; LV, left ventricular; LVH, left ventricular hypertrophy; MGH, Massachusetts General Hospital; Mod/Sev, moderate to severe; SHD, structural heart disease; XGBoost, extreme gradient boosting; YNHHS, Yale-New Haven Health System.

| Disease Label | Model Type | YNHHS | YNHHS | MGH | MGH | EUH | EUH |
|---|---|---|---|---|---|---|---|
| - | - | Concordance % | Cohen’s Kappa | Concordance % | Cohen’s Kappa | Concordance % | Cohen’s Kappa |
| LV Systolic Dysfunction | CNN | 84.30% | 0.633 (0.630-0.637) | 82.90% | 0.605 (0.603-0.608) | 81.90% | 0.625 (0.621-0.630) |
| Severe LVH | CNN | 84.20% | 0.670 (0.667-0.673) | 82.50% | 0.645 (0.643-0.647) | 83.60% | 0.672 (0.668-0.677) |
| Mod/Sev Valve Disease | CNN | 84.40% | 0.676 (0.673-0.678) | 82.20% | 0.631 (0.629-0.633) | 83.20% | 0.619 (0.613-0.625) |
| Mitral Regurgitation | CNN | 82.50% | 0.648 (0.645-0.651) | 80.40% | 0.604 (0.602-0.606) | 81.30% | 0.599 (0.594-0.604) |
| Aortic Stenosis | CNN | 84.10% | 0.681 (0.678-0.684) | 82.60% | 0.651 (0.649-0.653) | 82.30% | 0.640 (0.634-0.646) |
| Aortic Regurgitation | CNN | 82.70% | 0.615 (0.612-0.618) | 80.60% | 0.570 (0.568-0.573) | 82.10% | 0.560 (0.553-0.565) |
| Composite SHD | XGBoost | 89.10% | 0.779 (0.776-0.782) | 87.30% | 0.742 (0.740-0.745) | 86.80% | 0.716 (0.712-0.722) |

**Supplementary Table 5. Test-retest reliability of the image-based ensemble composite (Practical Screening using Ensemble Machine Learning Strategy for Structural Heart Disease) across real-world electrocardiogram image acquisition modalities and novel plotting formats.** Abbreviations: CCC, concordance correlation coefficient; CI, confidence intervals; ECG, electrocardiograms; EHR, electronic health record.

| | ECG Image Type | Number | CCC (95% CI) | Cohen's Kappa* (95% CI) | Concordance % |
|---|---|---|---|---|---|
| **Real-world Images** | **ECG Image Screenshots from EHR** | 100 | 0.973 (0.959, 0.982) | 0.796 (0.667, 0.918) | 90 |
| | **Smartphone Photos of ECGs on Computer Screen** | 100 | 0.967 (0.948, 0.983) | 0.837 (0.723, 0.940) | 92 |
| | **Smartphone Photos of ECG Printouts** | 100 | 0.956 (0.933, 0.974) | 0.779 (0.641, 0.899) | 89 |
| **Novel Plotted Formats** | **Novel Background Grid Color** | 10986 | 0.993 (0.993, 0.994) | 0.943 (0.937, 0.950) | 97.2 |
| | **Novel ECG Trace Color** | 10986 | 0.998 (0.998, 0.998) | 0.971 (0.966, 0.975) | 98.6 |
| | **Novel ECG Lead Layout** | 10986 | 0.963 (0.961, 0.965) | 0.862 (0.852, 0.871) | 93.3 |
| | **Novel Grid, Trace Colors & Lead Layout** | 10986 | 0.964 (0.962, 0.966) | 0.857 (0.846, 0.866) | 93.1 |

* Calculated at threshold for 90% sensitivity in internal testing

**Supplementary Table 6. Demographic and clinical characteristics of the predictive analysis cohort (paired electrocardiograms 30-90 days apart, individuals at risk for new-onset heart failure) across three health systems.** Abbreviations: DM, type-2 diabetes mellitus; ECG, electrocardiogram; HF, heart failure; HTN, hypertension; IQR, interquartile range.

| **Model** | **YNHHS** | **MGH** | **EUH** |
|---|---|---|---|
| **Number of Patients** | 39517 | 22940 | 3381 |
| **Age (years; Median [IQR])** | 62.3 [48.1,74.3] | 62.9 [51.0,71.6] | 65.6 [54.5,73.8] |
| **Female Sex (%)** | 20465 (51.8) | 11490 (50.1) | 1661 (49.1) |
| **Race/Ethnicity (%)** | **-** | **-** | **-** |
| **Non-Hispanic White** | 27076 (68.5) | 17019 (74.2) | 1970 (58.3) |
| **Non-Hispanic Black** | 5632 (14.3) | 2098 (9.1) | 1085 (32.1) |
| **Hispanic** | 4790 (12.1) | 2346 (10.2) | 12 (0.4) |
| **Non-Hispanic Asian** | 693 (1.8) | 695 (3.0) | 60 (1.8) |
| **Non-Hispanic Others** | **-** | 175 (0.8) | 199 (5.9) |
| **Unknown** | 1326 (3.4) | 607 (2.6) | 55 (1.6) |
| **Follow up (years; Median [IQR])** | 4.7 [2.1,7.5] | 3.2 [1.5,5.1] | 5.4 [1.8,6.4] |
| **AI-ECG Test Transitions** | **-** | **-** | **-** |
| **Positive-Positive** | 15523 (39.3) | 8204 (35.8) | 1763 (52.1) |
| **Negative-Positive** | 2174 (5.5) | 1589 (6.9) | 261 (7.7) |
| **Positive-Negative** | 4239 (10.7) | 1673 (7.3) | 260 (7.7) |
| **Negative-Negative** | 17581 (44.5) | 11474 (50.0) | 1097 (32.4) |
| **Absolute Delta Between ECGs (days; median [IQR])** | 4.7 [2.1,7.5] | 3.2 [1.5,5.1] | 5.4 [1.8,6.4] |
| **HTN (%)** | 25422 (64.3) | 10878 (47.4) | 356 (10.5) |
| **DM (%)** | 9707 (24.6) | 3866 (16.9) | 501 (14.8) |
| **Obesity (%)** | 8812 (22.3) | 3159 (13.8) | 309 (9.1) |
| **Outcome (New-onset HF) (%)** | 1876 (4.7) | 2099 (9.1) | 1122 (33.2) |

**Supplementary Table 7. Hazard ratios for new-onset heart failure by serial change in screen status of artificial intelligence-enhanced electrocardiography, in sequentially adjusted Cox proportional hazards models, across three health systems.** Abbreviations: CI, confidence intervals; DM, type-2 diabetes mellitus; HR, hazard ratio; HTN, hypertension.

| Model | AI-ECG Screen Status Changes | YNHHS: Hazard Ratio (95% CI) | MGH: Hazard Ratio (95% CI) | EUH: Hazard Ratio (95% CI) |
|---|---|---|---|---|
| Unadjusted Cox Model | Positive-Positive | 6.20 (5.46-7.04) | 4.58 (4.12-5.09) | 4.12 (3.48-4.88) |
| | Negative-Positive | 2.45 (1.91-3.14) | 2.44 (2.02-2.95) | 2.14 (1.62-2.84) |
| | Positive-Negative | 1.65 (1.33-2.05) | 2.31 (1.92-2.78) | 1.68 (1.25-2.26) |
| + Demographics (Age, Sex, Race/Ethnicity) | Positive-Positive | 3.47 (2.95-4.08) | 3.95 (3.46-4.52) | 4.79 (3.97-5.77) |
| | Negative-Positive | 1.89 (1.47-2.44) | 2.24 (1.85-2.72) | 2.32 (1.75-3.08) |
| | Positive-Negative | 1.46 (1.17-1.81) | 2.12 (1.75-2.57) | 1.82 (1.35-2.46) |
| + Comorbidities (HTN, DM, Obesity) | Positive-Positive | 3.18 (2.71-3.73) | 3.79 (3.33-4.33) | 4.64 (3.85-5.60) |
| | Negative-Positive | 1.67 (1.30-2.15) | 2.15 (1.77-2.61) | 2.37 (1.79-3.15) |
| | Positive-Negative | 1.30 (1.05-1.62) | 2.04 (1.69-2.47) | 1.83 (1.36-2.48) |
